## Supplementary materials for "A unifying model to explain high nirmatrelvir therapeutic efficacy against SARS-CoV-2, despite low post-exposure prophylaxis efficacy and frequent viral rebound"

Shadisadat Esmaeili *et al*.

**This file includes:**

Figs. S1 to S16

Tables S1 to S5

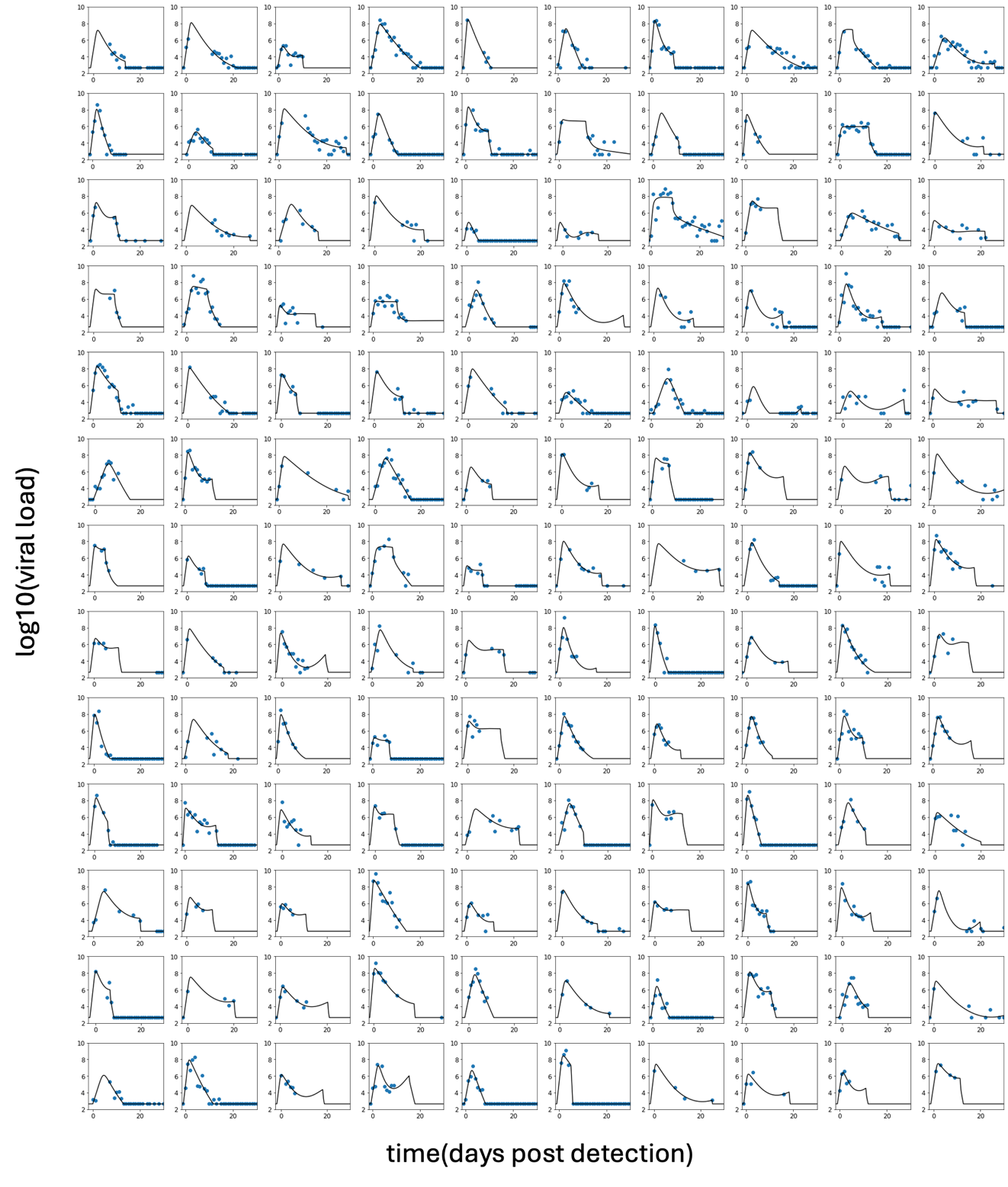

**Fig S1. Viral dynamics model recapitulation of untreated SARS-CoV-2 kinetics**. Viral dynamics model fit to diverse individual data. Included panels are 130 sample individuals from the NBA cohort with positive symptom status.

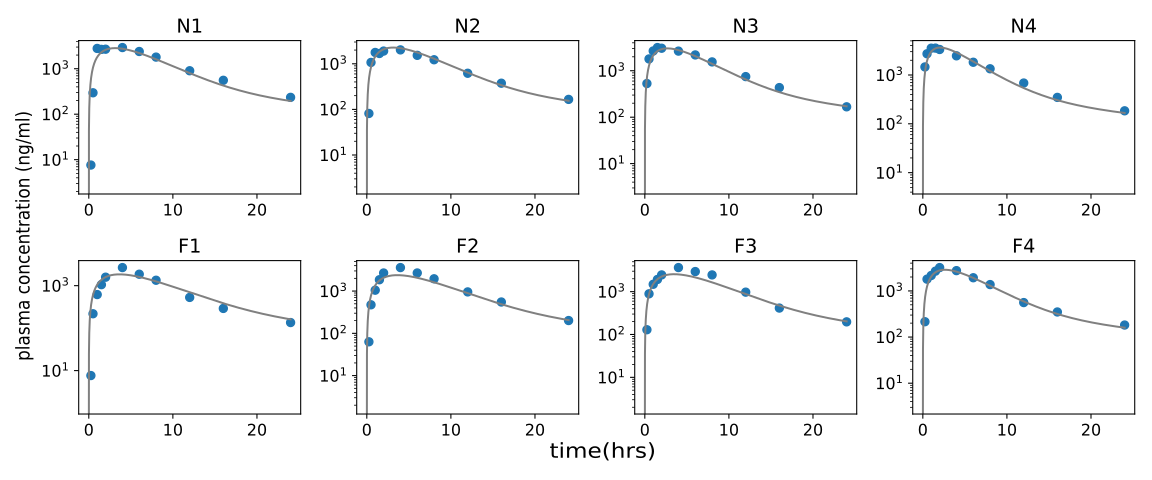

**Fig S2. Pharmacokinetic (PK) model recapitulation of plasma concentration of nirmatrelvir.** PK model fits individual nirmatrelvir/ritonavir plasma concentration data of 8 individuals (4 fed (F), 4 fasting (N)) with the 250 mg/100 mg dose of nirmatrelvir/ritonavir. Fed and fasting refer to individuals taking the drug under fed and fasting conditions, as explained in the original study by Singh et al (2022).

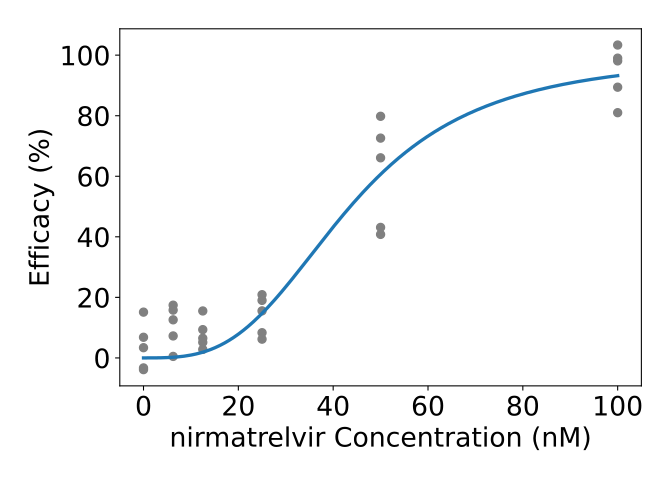

**Fig S3. Pharmacodynamic (PD) model fitting to mean antiviral efficacy as a function of drug concentration in vitro.** PD model fitting to mean efficacy of nirmatrelvir+efflux inhibitor was obtained from 5 replicates pooled from 2 in vitro infection experiments in Calu3 cells using the least square method.

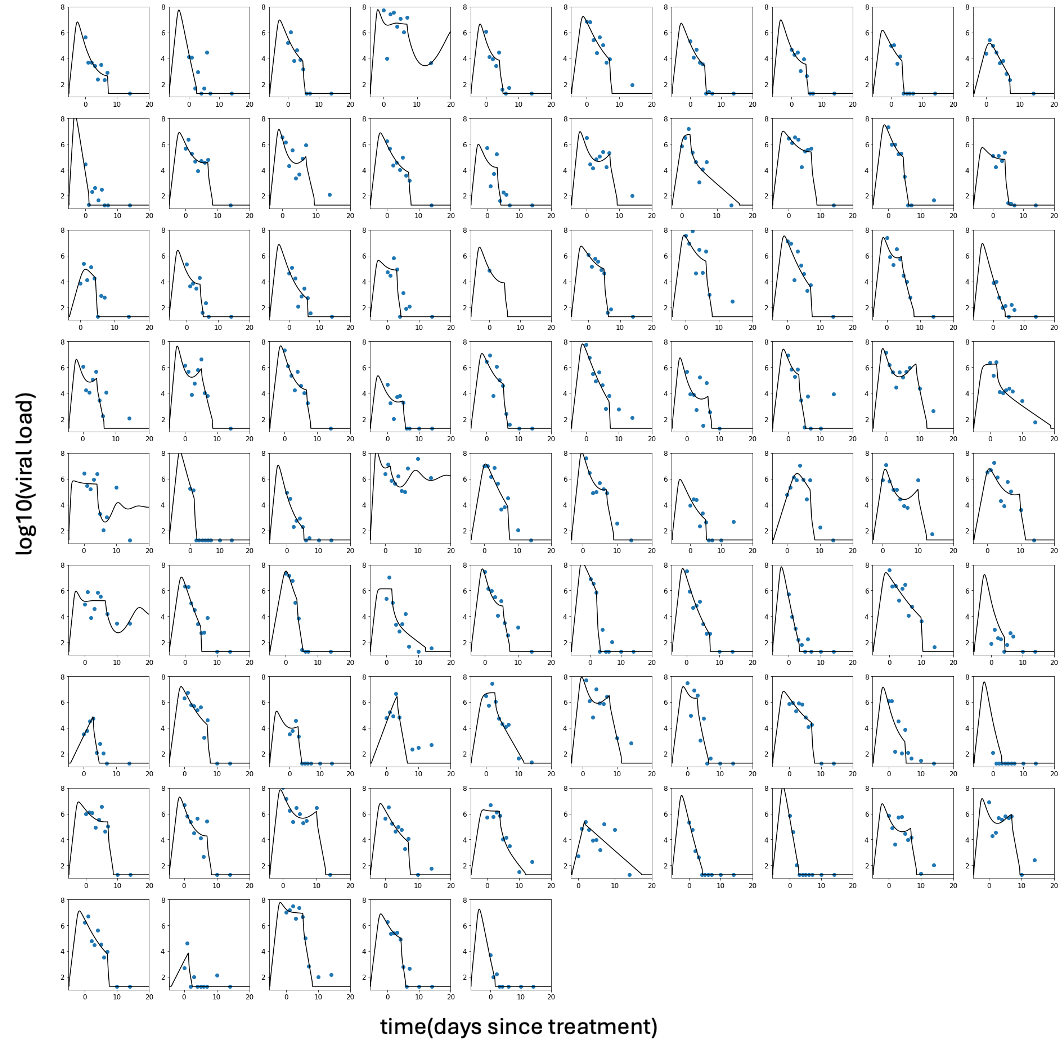

**Fig S4. Mathematical model recapitulation of the control arm of the PLATCOV trial**. Combined model fit to individual data.

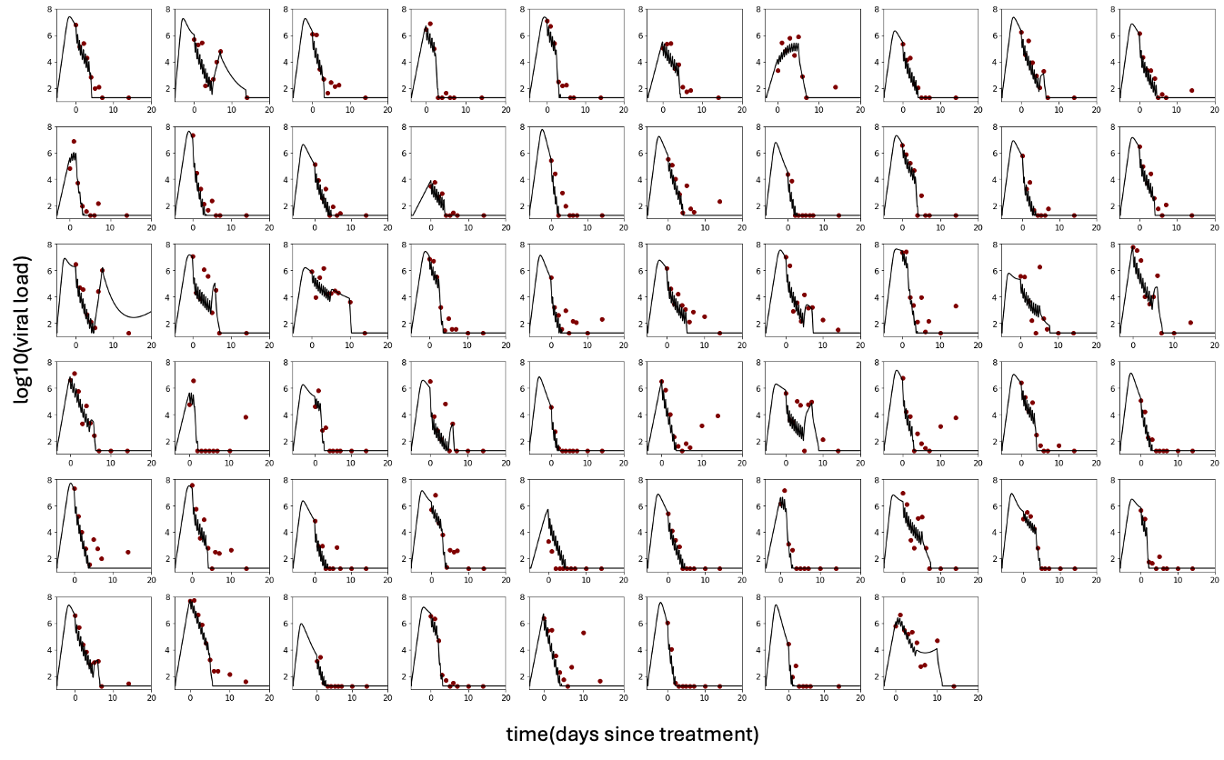

**Fig S5. Mathematical model recapitulation of the treatment arm of the PLATCOV trial**. Combined model fit to individual viral load data. PK and PD parameters were fixed for all individuals.

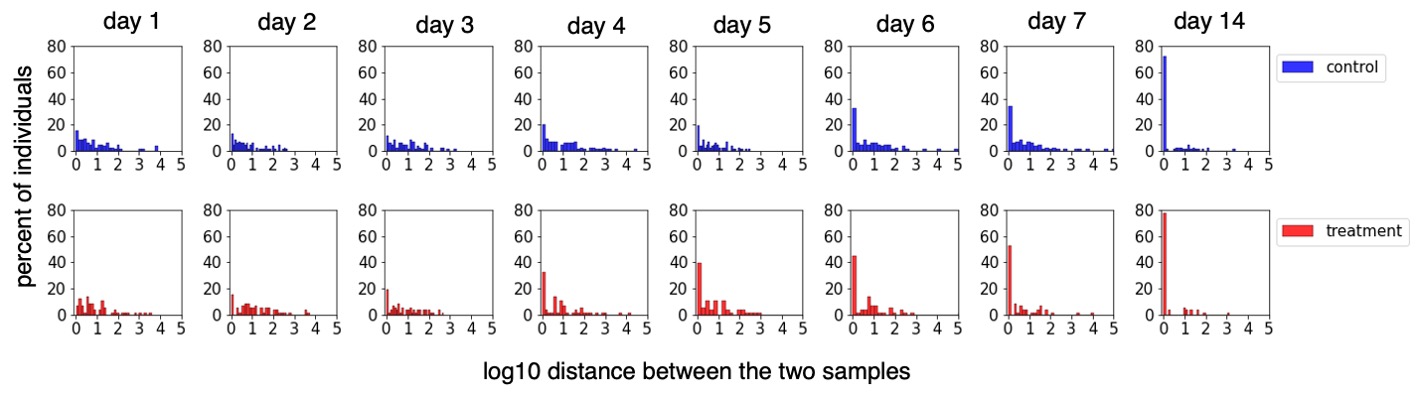

**Fig S6. Differences in SARS-CoV-2 viral loads from contemporaneous oral samples collected from each individual in the PLATCOV trial *(25)*.** The distribution of distance between the two samples collected from each participant’s left and right tonsil in the control arm (top row) and the treatment arm (bottom row). In certain instances, there is a substantial difference in viral loads between the two samples.

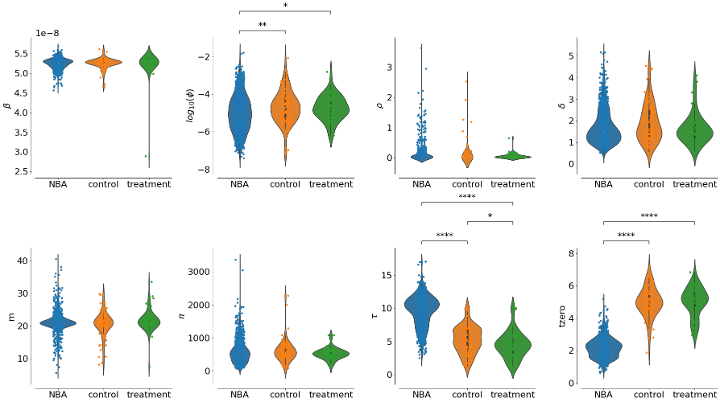

**Fig S7. Population parameter distributions for symptomatic Omicron-infected individuals of NBA cohort and the two arms of the PLATCOV trial.** In the PLATCOV trial $\tau$ and $t_{zero}$ are estimated relative to the time of the baseline measurement, while in the NBA cohort, they are estimated relative to the first positive test.

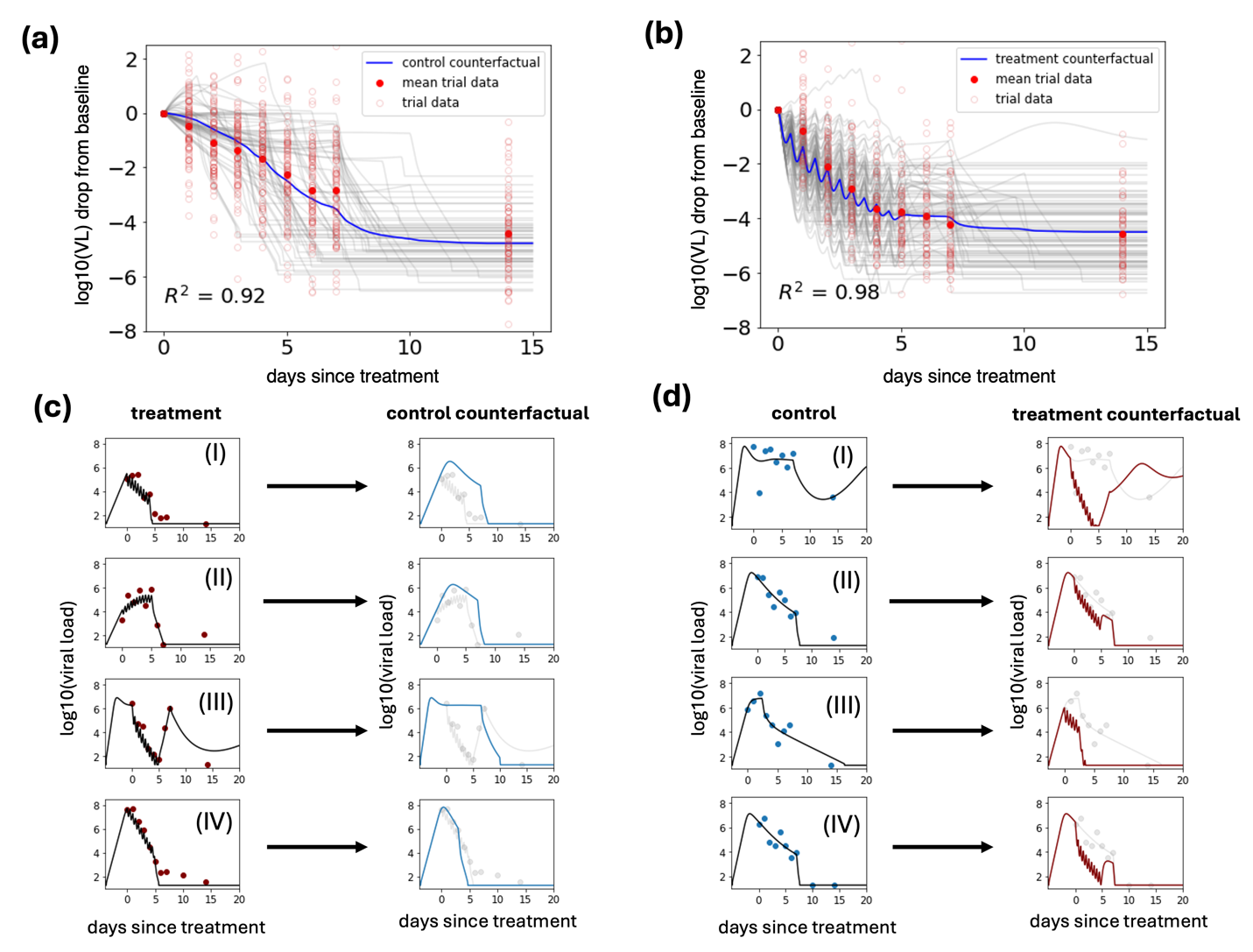

**Fig S8. Counterfactual simulations of the control and treatment arms of the PLATCOV trial recapitulate trial outcomes.** (a-b) individual simulated control counterfactual of the treatment arm **(a)** and the treatment counterfactual of the control arm **(b)** (in grey) and the mean viral load drop from the baseline (in blue) and the trial data. **(c-d)** sample model fits to the treatment arm and its control counterfactual simulation **(c)** and control arm and its treatment counterfactual simulation**(d)**.

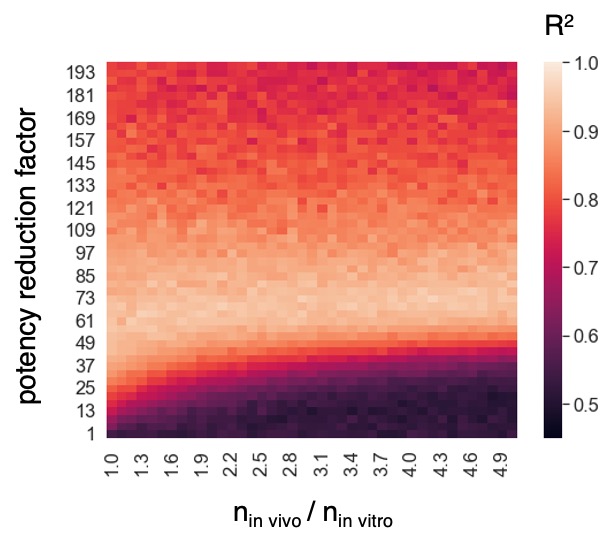

**Fig S9.** **The estimated potency reduction factor is independent of the in vivo Hill coefficient.** The R^2^ of the fit to the treatment arm of EPIC-HR for different prf values and Hill coefficients. The best fit with the highest R^2^ is obtained for prf ~ 60 and is independent of the Hill coefficient.

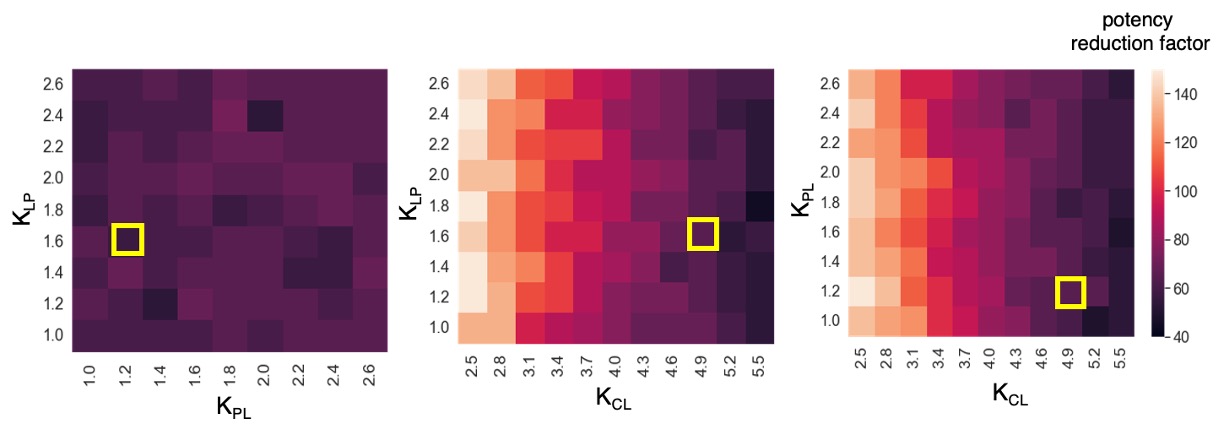

**Fig S10. Only the drug’s clearance rate affects the estimated potency reduction factor.** Potency reduction factor sensitivity to PK parameters fit to data from EPIC-HR. The yellow square in each panel marks the estimated values of the PK parameters by fitting the PK model to the trial data with single dose of 250mg/100mg nirmatrelvir/ritonavir.

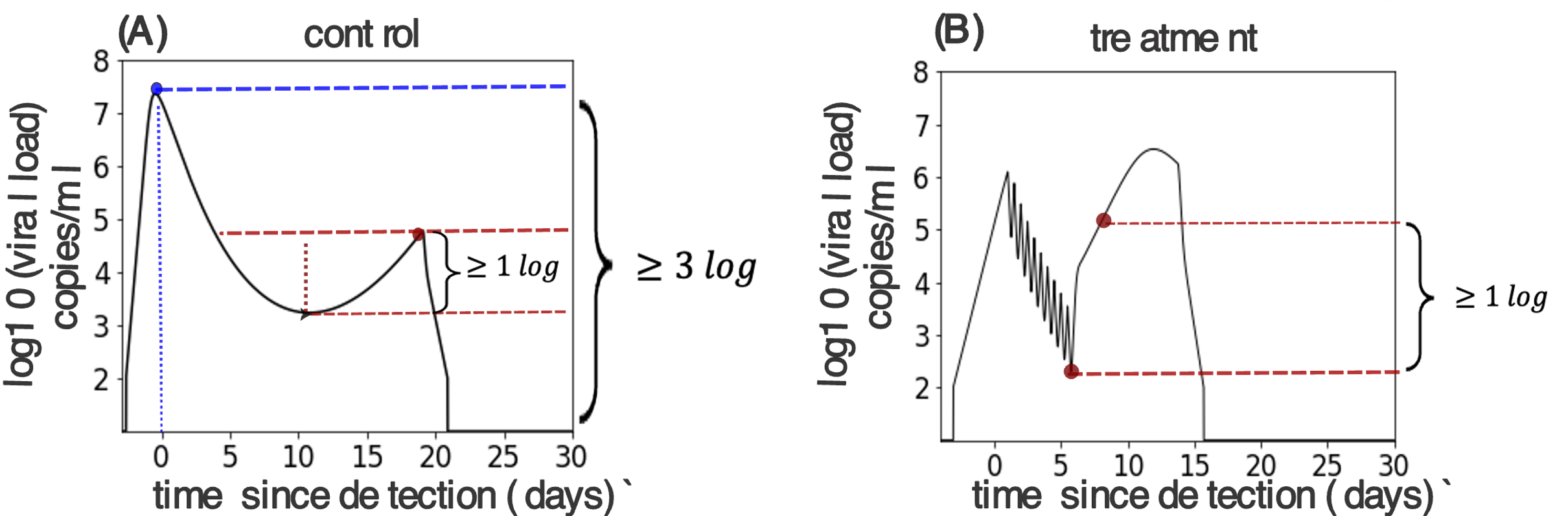

**Fig S11. Rebound definition in control and treatment arms. (A)** Rebound was flagged in the control arm if there were two peaks with a height greater than 3 logs and the second peak higher than the post-peak trough by at least 1 log. (B) Rebound in treatment arm was defined as increased viral load by at least 1 log any time after the end of treatment.

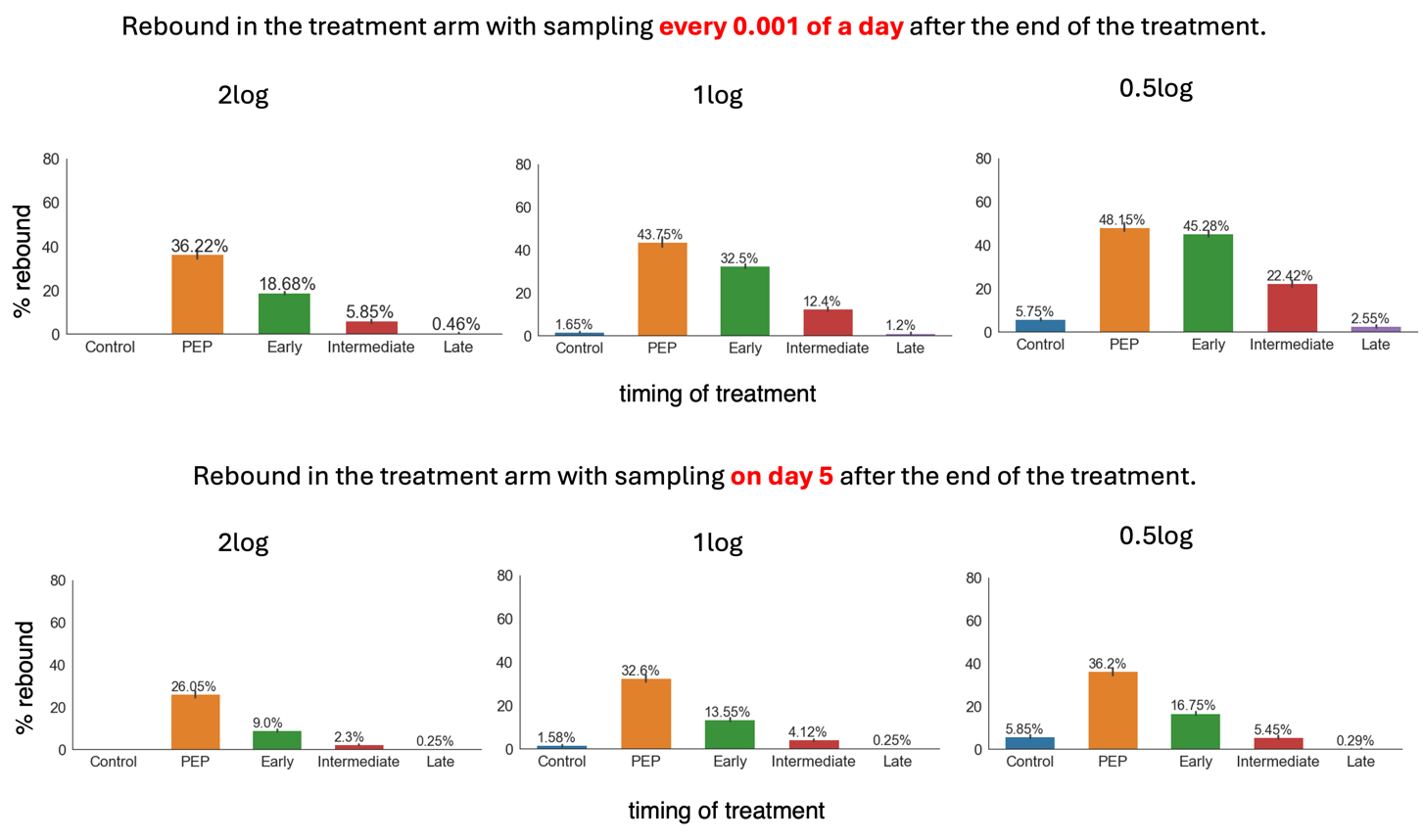

**Fig S12. Sensitivity of viral rebound detection to rebound definition and sampling frequency.** Top row: rebound probability in the treatment arm measured by sampling every 0.001 of a day after treatment and defining rebound as cases when viral load exceeds the viral load at the end of the treatment by 2 log, 1 log, or 0.5 log (left to right). Bottom row: rebound probability measured by sampling on day 5 after treatment ends with the same thresholds as the top row from left to right.

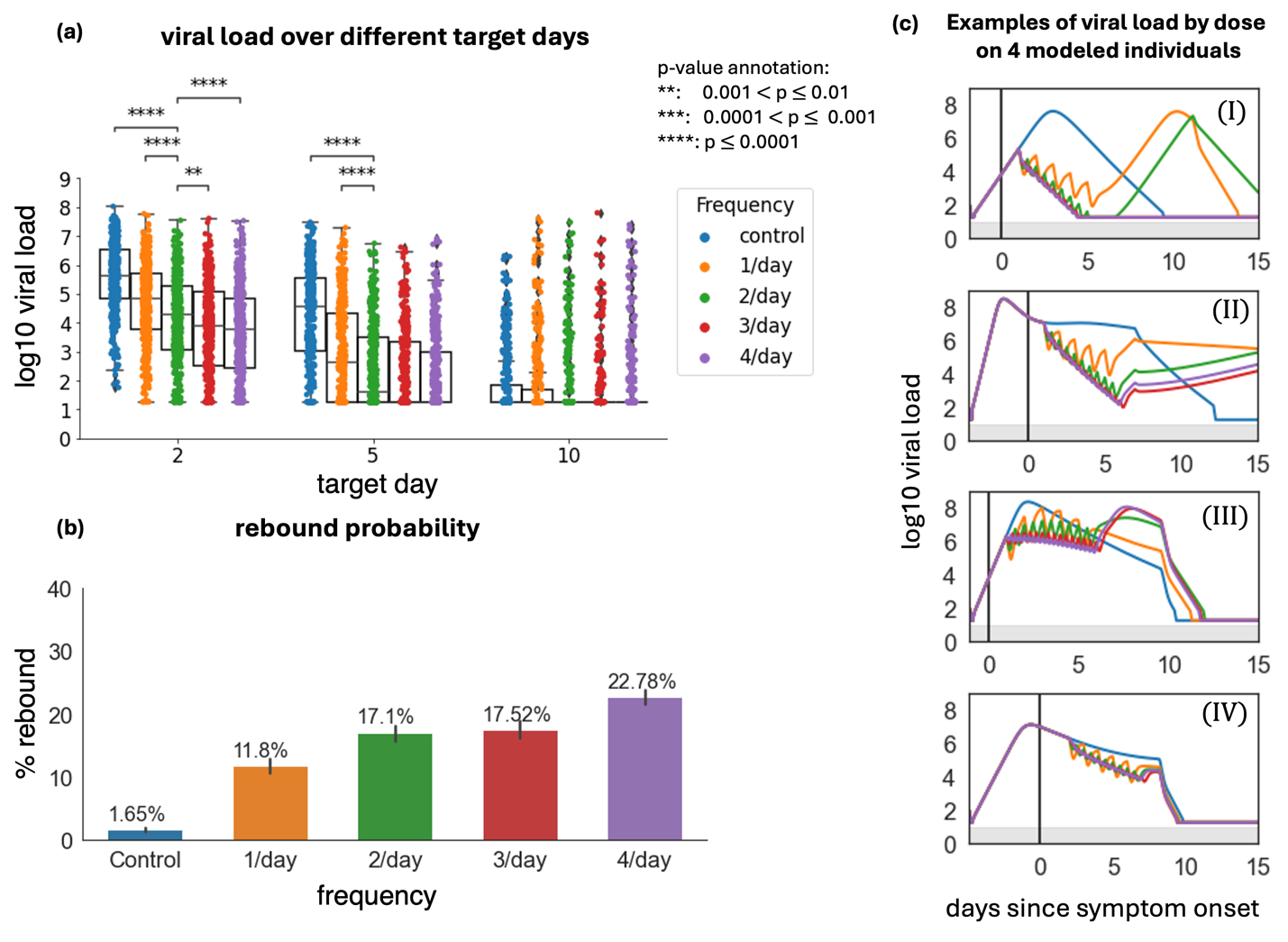

Fig. S13. Increasing nirmatrelvir dosing frequency lowers short term viral load but increases probability of viral rebound. In all simulations, 300 mg treatment starts within the first 3 days post-symptoms. (a) log10 viral load at days 2, 5, and 10 after the treatment start day with different dosing frequencies. p-values are obtained by performing two-sided Mann-Whitney U-test between 2/day group and others and only values <0.01 are shown. Viral loads are only reduced by higher dosing frequency at days 2 and 5, but not day 10. (b) The probability of rebound for different doses. The error bars on each column are 95% confidence interval. (c) Samples of viral load trajectories assuming different dosing frequency on 4 modeled individuals with equivalent timing of therapy and untreated viral kinetics.

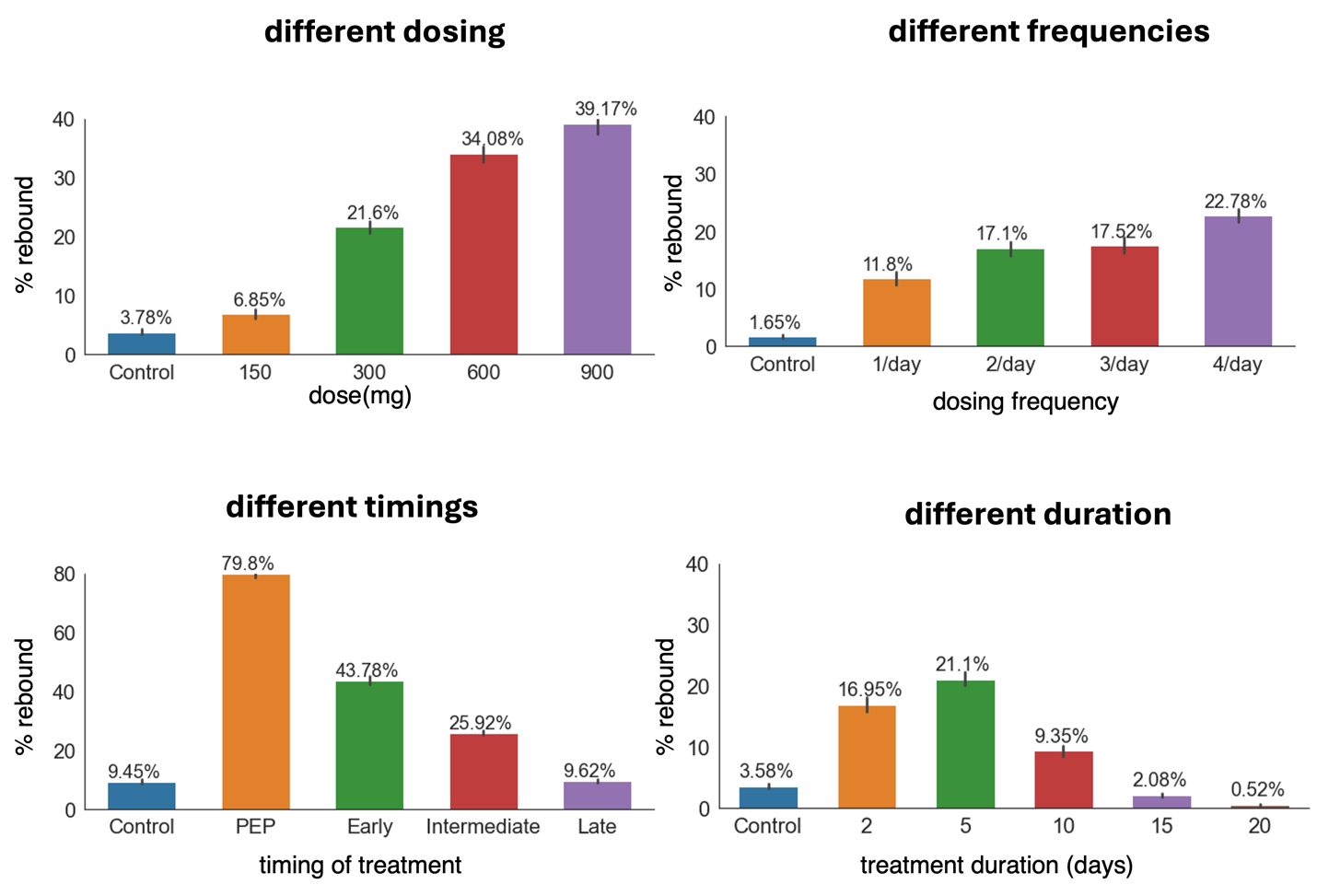

Fig. S14. Higher probabilities of viral rebound among unvaccinated individuals mostly infected with pre_Omicron VOC. In the control arm and all treatment scenarios the model predicts higher incidents of rebound in the unvaccinated pre_Omicron individuals. These results can be compared to Figs 4-7 and Fig S13 and show consistently higher rebound assuming equivalent conditions.

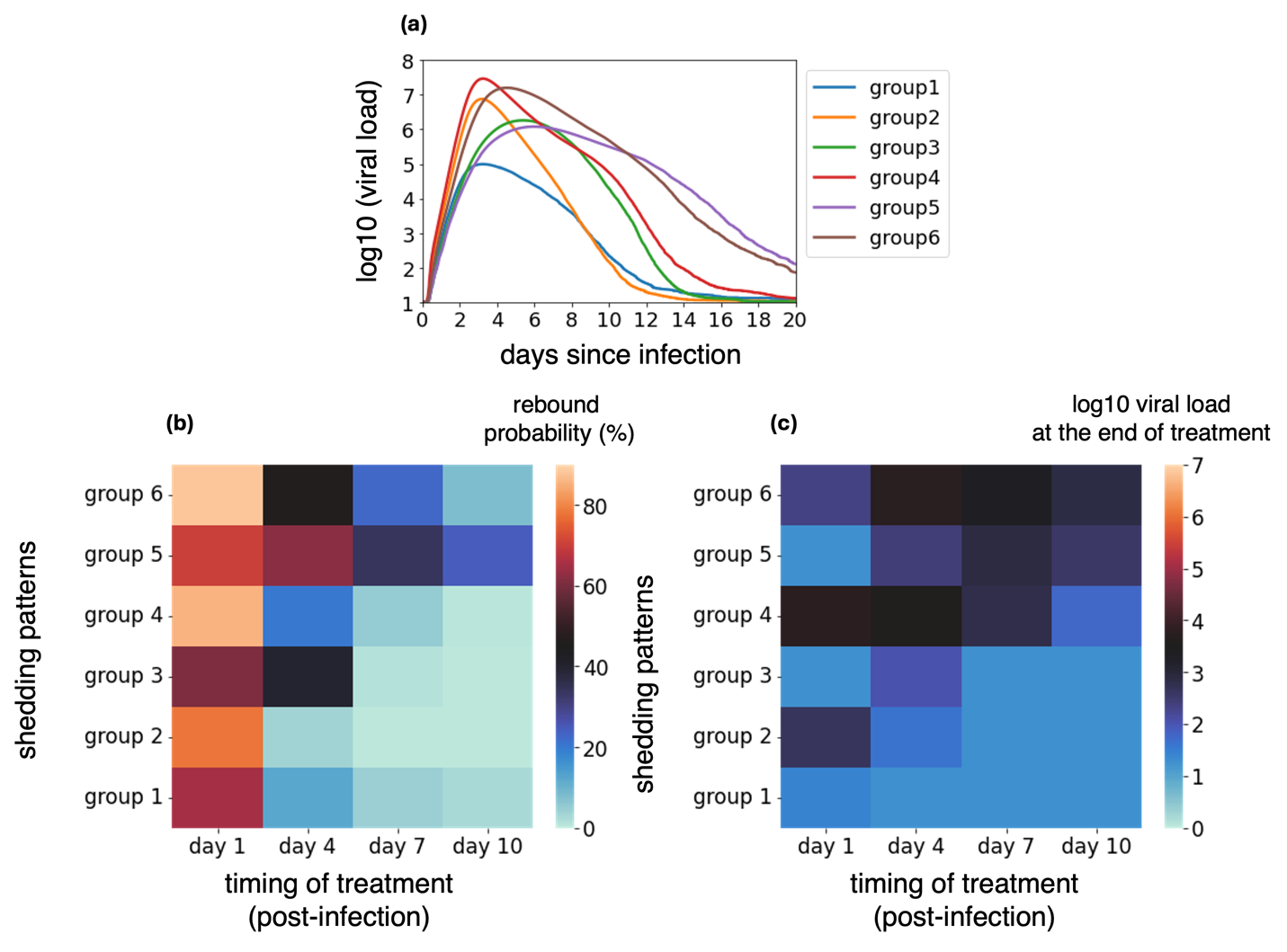

**Fig S15. Higher rebound probability in individuals with higher peak viral load and faster viral expansion after early treatment.** **(a)** Modeled mean viral load trajectories of six different groups off-treatment demonstrating differing peak viral load, time of viral peak, clearance rate, and time to clearance. Shedding pattern groups were from the NBA cohort identified in Owens et al. using k-means clustering. Higher group number indicates a higher viral area under the curve assuming untreated infection. **(b)** The probability of rebound and **(c)** the viral load at the end of treatment with different timing of treatment initiation.

| Parameter (unit) | Symbol | Population Mean | Standard error | Standard deviation of random effects | Distribution | Source |
| --- | --- | --- | --- | --- | --- | --- |
| viral infectivity  (log_10_ (RNAcopies/mL)^-1^ day^-1^) | log_10_$\beta$ | -7.31 | 3.17e-3 | 0.047 | normal | estimated |
| viral production rate  (log_10_ day^-1^ ) | log_10_$\pi$ | 2.74 | 9.7e-3 | 0.316 | normal | estimated |
| rate at which refractory cells revert to susceptible state  (log_10_ day^-1^ ) | log_10_$\rho$ | -1.62 | 3.33e-2 | 0.972 | normal | estimated |
| rate constant for conversion of target cells to a refractory state (log_10_ cell^-1^day^-1^ ) | log_10_$\phi$ | -5.29 | 6.52e-2 | 1.2 | normal | estimated |
| Deviation from $\log_{10} \phi$ for delta/other | $\beta_{\phi_{omicron}}$ | 0.324 | 7.38e-2 | -- | -- | -- |
| infected cell clearance rate (day^-1^ cells^-1^) | $\delta$ | 1.38 | 2.64e-2 | 0.565 | lognormal | estimated |
| onset of acquired immunity relative to detection (days) | $\tau$ | 15.9 | 0.453 | 0.461 | lognormal | estimated |
| Deviation from $\tau$ for delta/other | $\beta_{\tau_{omicron}}$ | -0.364 | 3.53e-2 | -- | -- | -- |
| Deviation from $\tau$ for unvaccinated/no record | $\beta_{\tau\_>1vax}$ | -0.141 | 3.16e-2 | -- | -- | -- |
| Increase in clearance rate of infected cells due to acquired immunity (day^-1^) | $m$ | 16.4 | 0.542 | 0.502 | lognormal | estimated |
| delay between infection in nasal tissue and detection (days) | $t_{0}$ | 2.08 | 2.8e-2 | 0.463 | logit[0,20] | estimated |
| initial viral inoculum (RNAcopies/mL) | $V_{0}$ | 97 | -- | -- | -- | fixed |
| viral clearance rate (day^-1^) | $\gamma$ | 15 | -- | -- | -- | Goyal et al. |
| mean eclipse phase duration (days^-1^) | $1/k$ | 1/4 | -- | -- | -- | Ke et al. |
| Initial number of susceptible cells | $S_{0}$ | $1 \times{10}^{7}$ | -- | -- | -- | Ortiz et al. |
| Initial number of refractory cells | $R_{0}$ | 0 | -- | -- | -- | -- |
| Initial number of productively infected cells | $I_{P,0}$ | 0 | -- | -- | -- | -- |
| Initial number of infected cells in eclipse phase | $I_{E,0}$ | 0 | -- | -- | -- | -- |

**Table S1. Population parameters of viral dynamics model*.*** Using the viral dynamic model, we estimated model parameters for 1510 infections in the NBA cohort. For the mixed-effect model, a constant error model was used, and the magnitude of measurement error was fixed at a = 0.25 log10 copies viral RNA/ml. Vaccine status (0 vs >1 dose) was set as a covariate on $\tau$, and the variance (other/delta vs omicron) was set as a covariate for $\tau$ and $\phi$. Also, linear dependencies between ($\pi, \phi, \delta)$ and ($\tau, \rho$) were set up in Monolix. The population parameters are recorded here and estimated individual parameters are available at https://github.com/sEsmaeili/Covid_Rebound.

| Parameter (unit) | Symbol | Population Mean | Standard deviation | Distribution |
| --- | --- | --- | --- | --- |
| Absorption rate  (day^-1^) | $\kappa_{a}$ | 9.98 | 0.37 | lognormal |
| Rate of transfer from plasma to lung  (day^-1^ ) | $\kappa_{PL}$ | 1.58 | 0.1 | lognormal |
| Rate of transfer from lung to plasma  (day^-1^ ) | $\kappa_{LP}$ | 1.22 | 0.1 | lognormal |
| Clearance rate (day^-1^) | $\kappa_{CL}$ | 4.96 | 0.03 | lognormal |
| Plasma Volume (ml) | $Vol$ | 41743 | 0.12 | lognormal |
| Error parameter | b | 0.34 |  |  |

**Table S2. Population parameters of pharmacokinetic model*.*** Using the two-compartmental PK model with oral administration of the drug, and proportional error model (estimated error parameter b=0.34), we estimated model parameters for 8 healthy individuals in the phase I randomized clinical trial by Singh et al. (2022) .The population parameters are recorded here and estimated individual parameters are available at https://github.com/sEsmaeili/Covid_Rebound.

|  | $\kappa_{a}$ | $\kappa_{LP}$ | $\kappa_{CL}$ | $\kappa_{PL}$ | $Vol$ |
| --- | --- | --- | --- | --- | --- |
| 250(Fed) | 8.81 | 1.28 | 4.77 | 1.72 | 42571.61 |
| 250(fasting) | 12.32 | 1.28 | 4.78 | 1.76 | 40379.71 |
| 750(Fed) | 8.49 | 1.29 | 4.78 | 1.92 | 54274.78 |

**Table S3. Population parameters of pharmacokinetic model for different doses*.***

Similar PK parameters were estimated for different doses by fitting the two-compartmental PK model with oral administration of the drug to population-level plasma concentrations at different doses. Fed and fasting refer to individuals who took the drug fed or fasting conditions as explained in the original study by Singh et al (2022).

| Parameter (unit) | Symbol | Mean | Standard Error |
| --- | --- | --- | --- |
| Maximum efficacy (%) | $E_{max}$ | 99.9 | 0.026 |
| Drug concentration to provide 50% efficacy ($nMol$ ) | IC_50_ | 43.6 | 3.2 |
| Hill coefficient | $n$ | 3.16 | 0.42 |

**Table S4. Pharmacodynamic parameters*.*** Using the least square method, we estimated the PD parameters by fitting the Hill equation to the in vitro data.

| Parameter (unit) | Symbol | Population Mean | Standard error | Std dev. of random effects | Distribution | Source |
| --- | --- | --- | --- | --- | --- | --- |
| viral infectivity  (log_10_ (RNAcopies/mL)^-1^ day^-1^) | log_10_$\beta$ | -7.28 | 4.23e-3 | 5.66e-2 | normal | estimated |
| viral production rate  (log_10_ day^-1^ ) | log_10_$\pi$ | 2.72 | 1.12e-2 | 0.311 | normal | estimated |
| rate at which refractory cells revert to susceptible state  (log_10_ day^-1^ ) | log_10_$\rho$ | -1.57 | 3.87e-2 | 0.932 | normal | estimated |
| rate constant for conversion of target cells to a refractory state (log_10_ cell^-1^day^-1^ ) | log_10_$\phi$ | -5.01 | 4.05e-2 | 1.18 | normal | estimated |
| infected cell clearance rate (day^-1^ cells^-1^) | $\delta$ | 1.49 | 3.36e-2 | 0.556 | lognormal | estimated |
| onset of acquired immunity relative to detection (days) | $\tau$ | 10.8 | 0.405 | 0.473 | lognormal | estimated |
| Deviation from $\tau$ for unvaccinated/no record | $\beta_{\tau\_>1vax}$ | -0.057 | -- | -- | -- | -- |
| Deviation from $\tau$ for NBA Omicron Individuals | $\beta_{\tau\_PLATCOV}$ | -0.831 | -- | -- | -- | -- |
| Increase in clearance rate of infected cells due to acquired immunity (day^-1^) | $m$ | 20.9 | 0.953 | 0.548 | lognormal | estimated |
| time of infection relative to first detection (days) | $t_{0}$ | 2.13 | 3.46e-2 | 0.456 | logit[0,20] | estimated |
| Deviation from $t_{0}$ for NBA | $\beta_{t_{0},PLATCOV}$ | 1.05 | 6.32e-2 | -- | -- | -- |
| Potency reduction factor for PLATCOV treatment arm | prf | 40.3 | 5.48 | 0.826 | lognormal | estimated |
| initial viral inoculum (RNAcopies/mL) | $V_{0}$ | 97 |  | -- | -- | fixed |
| viral clearance rate (day^-1^) | $\gamma$ | 15 |  | -- | -- | Goyal et al. |
| mean eclipse phase duration (days^-1^) | $1/k$ | 1/4 |  | -- | -- | Ke et al. |
| Initial number of susceptible cells | $S_{0}$ | $1 \times{10}^{7}$ |  | -- | -- | Ortiz et al. |
| Initial number of refractory cells | $R_{0}$ | 0 |  | -- | -- | -- |
| Initial number of productively infected cells | $I_{P,0}$ | 0 |  | -- | -- | -- |
| Initial number of infected cells in eclipse phase | $I_{E,0}$ | 0 |  | -- | -- | -- |

**Table S5. Population parameters of combined viral dynamics + PKPD model fit to PLATCOV + NBA Omicron data*.*** Using the viral dynamic+PKPD model we estimated model parameters for Omicron infections in the NBA cohort and control and treatment arms of PLATCOV trial. For the mixed-effect model, a constant error model was used, and the magnitude of measurement error was fixed at a = 0.25 log10 copies viral RNA/ml. Vaccine status (0 vs >1 dose) was set as a covariate on $\tau$, and the cohort (NBA vs PLATCOV) was set as a covariate for $\tau$ and $t_{0}$. Also, linear dependencies between ($\pi, \phi, \delta)$ and ($\tau, \rho$) were set up in Monolix. The population parameters are recorded here and estimated individual parameters are available at https://github.com/sEsmaeili/Covid_Rebound.
